# Syndromic surveillance for mpox during a large religious mass gathering amid an ongoing outbreak, Uganda, 2025: a cross-sectional study

**DOI:** 10.64898/2026.08.12.26360261

**Authors:** Justine Wobusobozi, Richard Migisha, Martha Dorcas Nalweyiso, Winfred Nakaweesi, Aminah Namwabira, Kyomugisha Denise Aman, Benard Lubwama, Lilian Bulage, Benon Kwesiga, Alex Riolexus Ario

## Abstract

**Background:** Large religious mass gatherings can facilitate infectious disease transmission through crowding, prolonged close contact, shared facilities, and extensive population movement. The 2025 Uganda Martyrs’ Day commemoration occurred during an ongoing national mpox outbreak. We describe the implementation of syndromic surveillance for mpox during a large religious mass gathering and the yield of mpox-compatible illness detected among pilgrims.

**Methods:** We conducted a cross-sectional symptom-screening survey and retrospectively reviewed emergency medical services (EMS) records from the Namugongo Catholic and Protestant shrines during 29 May–5 June 2025. Pilgrims aged ≥18 years were selected through systematic sampling at entrance gates and random sampling within demarcated zones. Mpox-compatible illness was defined as acute fever (≥38.5°C) with rash and at least one of headache, lymphadenopathy, back pain, myalgia, or profound weakness. We summarized participant characteristics, reported symptoms, mpox-compatible illness, and provisional diagnoses among pilgrims seeking care.

**Results:** Of 1,523 pilgrims approached, 1,299 participated (85.3% response rate); median age was 35 years (interquartile range: 25–49), and 752 (57.9%) were male. Among actively screened pilgrims, 73 (5.6%) reported ≥2 mpox-related symptoms, and seven (0.5%) met the mpox-compatible illness definition. Among 5,216 pilgrims who sought care, 3,415 (65.5%) had documented presenting symptoms; of these, 628 (18.4%) had mpox-related syndromic presentations. The most common provisional diagnoses were peptic ulcer disease (n=563; 10.8%), respiratory tract infection (n=482; 9.2%), malaria (n=190; 3.6%), urinary tract infection (n=96; 1.8%), and gastritis (n=79; 1.5%).

**Conclusion:** Active symptom screening suggested a low prevalence of mpox-compatible illness in the general pilgrim population, whereas EMS records identified a larger syndromic signal among pilgrims who sought care. Combining both approaches with standardized documentation, clinical triage, laboratory confirmation, and follow-up could improve detection during mass gatherings.

## Background

Mass gatherings create conditions that can amplify infectious disease transmission because large numbers of people converge in a limited space, often with prolonged close contact, shared facilities, and substantial population movement before and after the event (1–3). These risks are especially important for religious gatherings, where crowding, communal activities, and cross-border attendance may increase opportunities for respiratory, gastrointestinal, skin, and close-contact infections to spread (4, 5). Syndromic surveillance offers a practical approach for monitoring such events because it uses clinical signs and symptoms to detect unusual illness patterns before laboratory confirmation is available (5, 6). This approach has been successfully implemented at various large-scale religious, sporting, and ceremonial events globally (6, 7). The Uganda Martyrs’ Day commemoration in Namugongo is one of Africa’s largest annual religious mass gatherings, attracting an estimated 2–3 million pilgrims from Uganda and neighbouring countries, making it an important setting for event-based public health surveillance (8).

The 2025 Uganda Martyrs’ Day commemoration occurred against the backdrop of an ongoing national mpox outbreak. By May–June 2025, thousands of confirmed cases had been reported across multiple districts, indicating widespread transmission(9). The event posed a particular surveillance challenge because mpox can spread through close physical contact, while early symptoms may be non-specific before characteristic skin lesions are recognized (10, 11). Syndromic surveillance conducted during the 2022 commemoration had described common acute illnesses and implementation gaps, but it preceded the mpox outbreak (8). Thus, evidence was limited on the detection of mpox-compatible illness during a large religious gathering held amid active community transmission.

Large mass gatherings held during ongoing outbreaks require surveillance systems that can rapidly identify possible cases while minimizing false alerts from common, non-specific illnesses. The 2025 Uganda Martyrs’ Day commemoration provided an opportunity to examine this challenge during active mpox transmission. We described the implementation of syndromic surveillance for mpox during a large religious mass gathering and assess its utility in detecting mpox-compatible illness among pilgrims attending the commemoration in Namugongo, Uganda.

## Methods

### Study design and setting

We conducted a cross-sectional study that combined a symptom screening survey with a retrospective review of emergency medical services (EMS) records. The study was carried out at the Namugongo Catholic and Protestant shrines in Namugongo Division, Wakiso District, Uganda (Figure 1), from 29 May to 5 June 2025, during the annual Uganda Martyrs’ Day commemoration.

**Figure 1.**
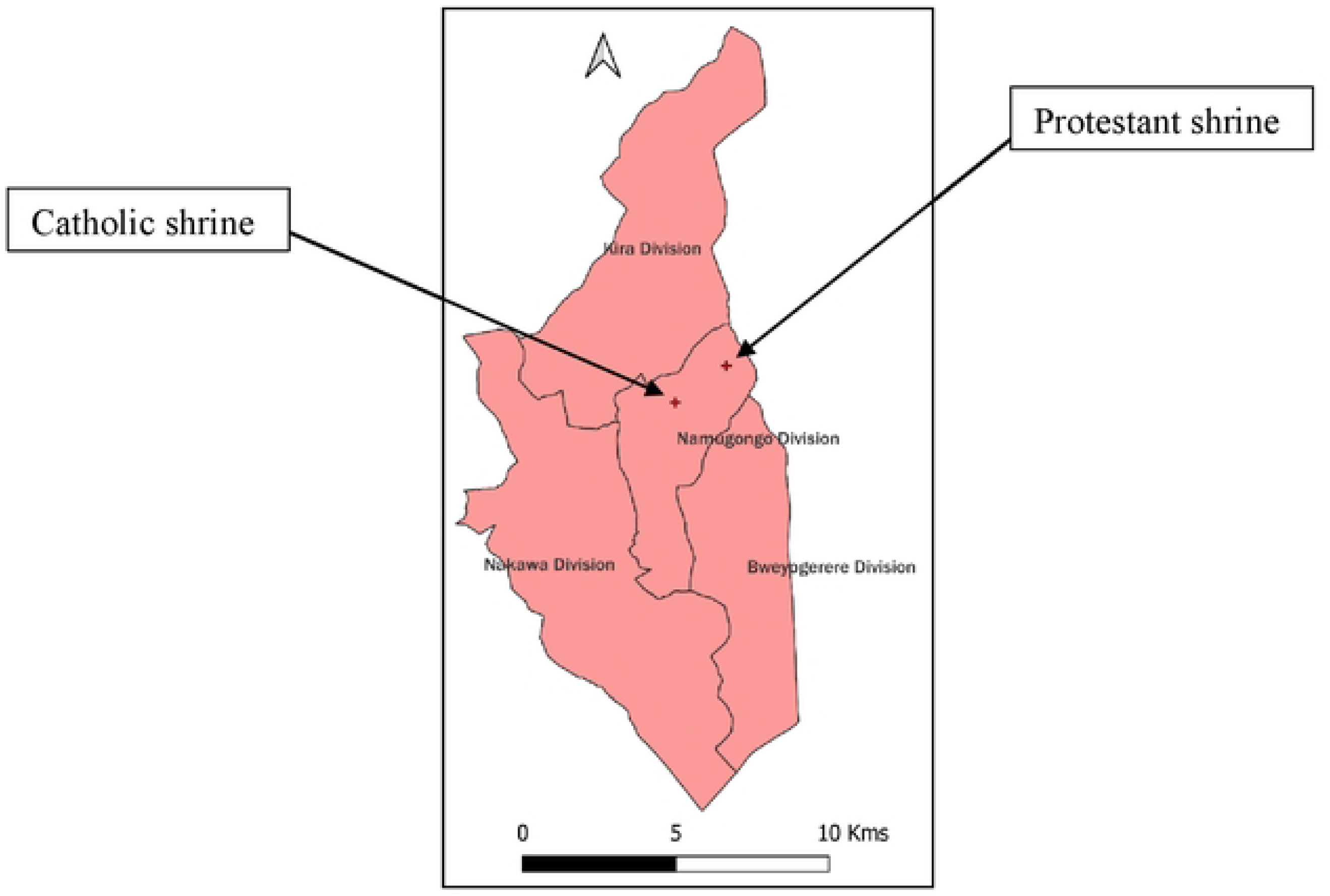
Location of the Namugongo Catholic and Protestant shrines in Wakiso District, Uganda, where syndromic surveillance was conducted during the 2025 Uganda Martyrs’ Day commemoration.

Pilgrims typically begin arriving at the Namugongo Catholic and Protestant shrines more than one week before the main commemoration on 3 June. Long-distance foot pilgrims from across Uganda and neighbouring countries often commence their journeys in mid-May and begin camping on-site from late May onwards. Attendance peaks on the commemoration day (3 June), after which most pilgrims gradually depart in the days immediately following the event. The study period from 29 May to 5 June 2025 therefore captured the late build-up phase, peak attendance, and immediate post-event phase of this mass gathering.

### Study population and exclusion criteria

The study population included pilgrims attending the commemoration. For the survey component, eligible participants were pilgrims aged ≥18 years who gave verbal informed consent. We excluded individuals unable to provide consent and non-pilgrims. For the EMS records review, we included all records of pilgrims who sought care at the designated on-site medical sites during the study period.

### Sample size and sampling strategy

The sample size for the survey was calculated using the Kish-Leslie formula for estimating a single proportion in a cross-sectional study, adjusted for design effect:

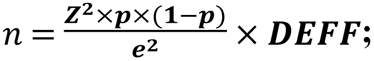 where Z=1.96 (for 95% confidence interval), p=0.05 (expected prevalence of mpox-compatible symptoms), e=0.02 (margin of error), and DEFF=1.5 (design effect for clustering). This yielded a target sample size of approximately 1,500 participants.

We enrolled participants using systematic sampling at the main entrance gates (every 10th pilgrim in line during peak entry hours, when thousands of pilgrims were expected to arrive daily based on historical attendance) and random selection within demarcated zonal areas (prayer zones and camping areas). Allocation was proportional to expected attendance: approximately 85% from the Catholic shrine and 15% from the Protestant shrine based on historical attendance patterns reported in prior surveillance (8). We enrolled 1,299 pilgrims. The shortfall from the target of 1,500 was due to extremely high crowd density that limited interviewer mobility, variable pilgrim flow rates on certain days, and higher-than-expected refusal rates in some zones during peak hours. The EMS component included all available records (n = 5,216) as a complete census.

### Study variables

Mpox-compatible illness was defined as acute onset of fever (≥38.5°C) followed by rash and at least one of the following: headache, lymphadenopathy, back pain, myalgia, or profound weakness (12). Other mpox-related syndromes were assessed using standard operational definitions (Table 1).

**Table 1:** Operational definitions of mpox and mpox-related syndromes.

| Syndrome | Definition |
| --- | --- |
| Suspected mpox | Acute onset of fever ($>38.5^{\circ}\text{C}$) and rash (maculopapular, vesicular, or pustular) AND at least one of: headache, lymphadenopathy, back pain, myalgia, or profound weakness. |
| Respiratory Tract Infection | Cough, sore throat, or difficulty breathing with or without fever. |
| Malaria | Fever, with positive malaria RDT, chills or headaches |
| Cellulitis/ Skin infection | Localized skin redness, swelling, warmth and pain suggestive of secondary bacterial infection of skin lesions |
| Pharyngitis | Sore throat, throat pain, or inflammation of the pharynx with or without fever |
| Conjunctivitis | Redness, itching, discharge or inflammation of one or both eyes |

*Adapted from World Health Organization mpox surveillance and case definitions (12) and* *Uganda Ministry of Health Integrated Disease Surveillance and Response (IDSR) technical* *guidelines (13).*

### Data collection

Surveillance consisted of two complementary components. Active symptom screening was conducted to identify pilgrims with possible mpox-compatible illness irrespective of healthcare seeking, while EMS surveillance captured syndromic presentations among pilgrims who sought medical care at on-site health facilities.

The survey was conducted among pilgrims at the Catholic and Protestant shrines from 29 May to 5 June 2025. The data collection tool was developed in KoboCollect based on signs and symptoms of mpox-compatible illness adapted from the World Health Organization mpox Case Investigation Form (14). The tool collected information on socio-demographic characteristics and self-reported mpox-compatible signs and symptoms. Trained surveillance officers conducted interviews anonymously, lasted 5-10 minutes and data were captured electronically using Kobo Collect software.

We also conducted records review of the on-site emergency medical records provided at the catholic and protestant shrines from 29 May to 5 June, 2025. We extracted all the available data of pilgrims who sought medical care from Health Management Information System registers for review including age, sex, district of residence, signs and symptoms or provisional diagnosis.

### Data management and statistical analysis

Survey data were downloaded from the KoboCollect server and cleaned in Excel. Active screening and EMS datasets were analyzed separately because they represented distinct surveillance populations. Descriptive statistics were used to summarize participant characteristics, reported symptoms, mpox-compatible illness, and provisional diagnoses. Continuous variables were summarized using medians and interquartile ranges, and categorical variables using frequencies and percentages. Analyses were performed using Epi Info 7 (CDC Atlanta, USA). No inferential statistics were performed because of the small number of mpox-compatible cases identified in the survey.

### Ethical consideration

This investigation was conducted in response to an annually commemorated mass gathering and was therefore determined to be non-research. In accordance with the Council for International Organizations of Medical Sciences (CIOMS) International Guidelines for Ethical Review of Epidemiological Studies (1991) and applicable CDC policies for emergency outbreak investigations, the activity was conducted consistent with applicable federal law and CDC policy (see, e.g., 45 C.F.R. part 46; 21 C.F.R. part 56; 42 U.S.C. §241[d]; 5 U.S.C. §552a; 44 U.S.C. §3501 et seq.).The Uganda National Institute of Public Health (UNIPH), under the Ministry of Health (MoH) approved the study protocols. The MoH gave authority and directive to conduct syndromic surveillance for mpox during this religious event. All methods were performed in accordance with approval and administrative clearance without any ethical breach.

Verbal consent in English and the local language was sought from pilgrims before participation in the survey. Systematically sampled participants were informed about the rationale of the survey and that their participation was voluntary without any negative consequences in case they refused. In accordance with the approved study protocol, there was a question in the screening section of the survey questionnaire inquiring as to whether the systematically sampled participant verbally consented; and for those who did not, they were skipped to the next 10th pilgrim in the line at main entrance gates. Pilgrims were assigned unique identifiers instead of using their names to protect the confidentiality of the respondents. Administrative clearance to extract patient data from Health Management Information System registers was obtained from the Ministry of Health. All methods were performed in accordance with the approval and administrative clearance

## Results

### Characteristics of pilgrims participating in active symptom screening

Of 1,523 pilgrims approached, 1,299 participated in the active symptom-screening survey, corresponding to a response rate of 85.3%. Age was recorded for 1,296 participants; the median age was 35 years (interquartile range [IQR]: 25–49 years). Overall, 752 (57.9%) participants were male, 1,261 (97.1%) were residents of Uganda, and 1,191 (91.7%) were recruited from the Catholic shrine (Table 2)

**Table 2:** Characteristics of pilgrims who participated in the survey during the Uganda Martyrs’ commemoration mass gathering, May 29–June 5, 2025.

| Characteristics | Frequency (n = 1,299) | Percentage (%) |
| --- | --- | --- |
| **Age\*** (n=1,296, due to 3 missing) |  |  |
| 18–29 years | 621 | 48 |
| 30–39 years | 233 | 18 |
| 40–49 years | 192 | 15 |
| ≥50 years | 250 | 19 |
| **Sex** |  |  |
| Male | 752 | 58 |
| Female | 547 | 42 |
| **Country of residence** |  |  |
| Uganda | 1,261 | 97 |
| Kenya | 28 | 2.2 |
| South Sudan | 1 | 0.1 |
| Rwanda | 1 | 0.1 |
| Democratic Republic of Congo | 1 | 0.1 |
| **Religious site visited** |  |  |
| Catholic shrine | 1,191 | 92 |
| Protestant shrine | 108 | 8 |

*n, number of participants; %, percentage; \*Median age (IQR): 35 (25 – 49)*

### Characteristics of pilgrims seeking medical care

During the surveillance period, 5,216 pilgrims sought care through on-site emergency medical services. Their median age was 38 years (IQR: 25–51 years); 3,548 (68.0%) were female and 1,668 (32.0%) were male. Pilgrims aged 18–29 years accounted for 1,560 (29.9%) care-seekers, while 1,521 (29.2%) were aged ≥50 years (Table 3).

**Table 3:** Characteristics of pilgrims who sought medical care from the medical tents during the Uganda Martyrs’ commemoration mass gathering, May 29 – June 5, 2025.

| Characteristics | Frequency (n = 5,216) | Percentage (%) |
| --- | --- | --- |
| **Age\*** |  |  |
| 18–29 years | 1,560 | 30 |
| 30–39 years | 1,051 | 20 |
| 40–49 years | 1,084 | 21 |
| $\geq 50$ years | 1,521 | 29 |
| **Sex** |  |  |
| Male | 1,668 | 32 |
| Female | 3,548 | 68 |

*n, number of participants; %, percentage; \*Median age (IQR): 38 (25 – 51)*

### Mpox-compatible illness identified through active screening

Among the 1,299 actively screened pilgrims, 496 (38.2%) reported at least one symptom included in the mpox screening algorithm, and 73 (5.6%) reported two or more such symptoms. Seven participants (0.5%) met the predefined mpox-compatible illness definition of acute fever with rash and at least one additional specified symptom (Figure 2).

**Figure 2.**
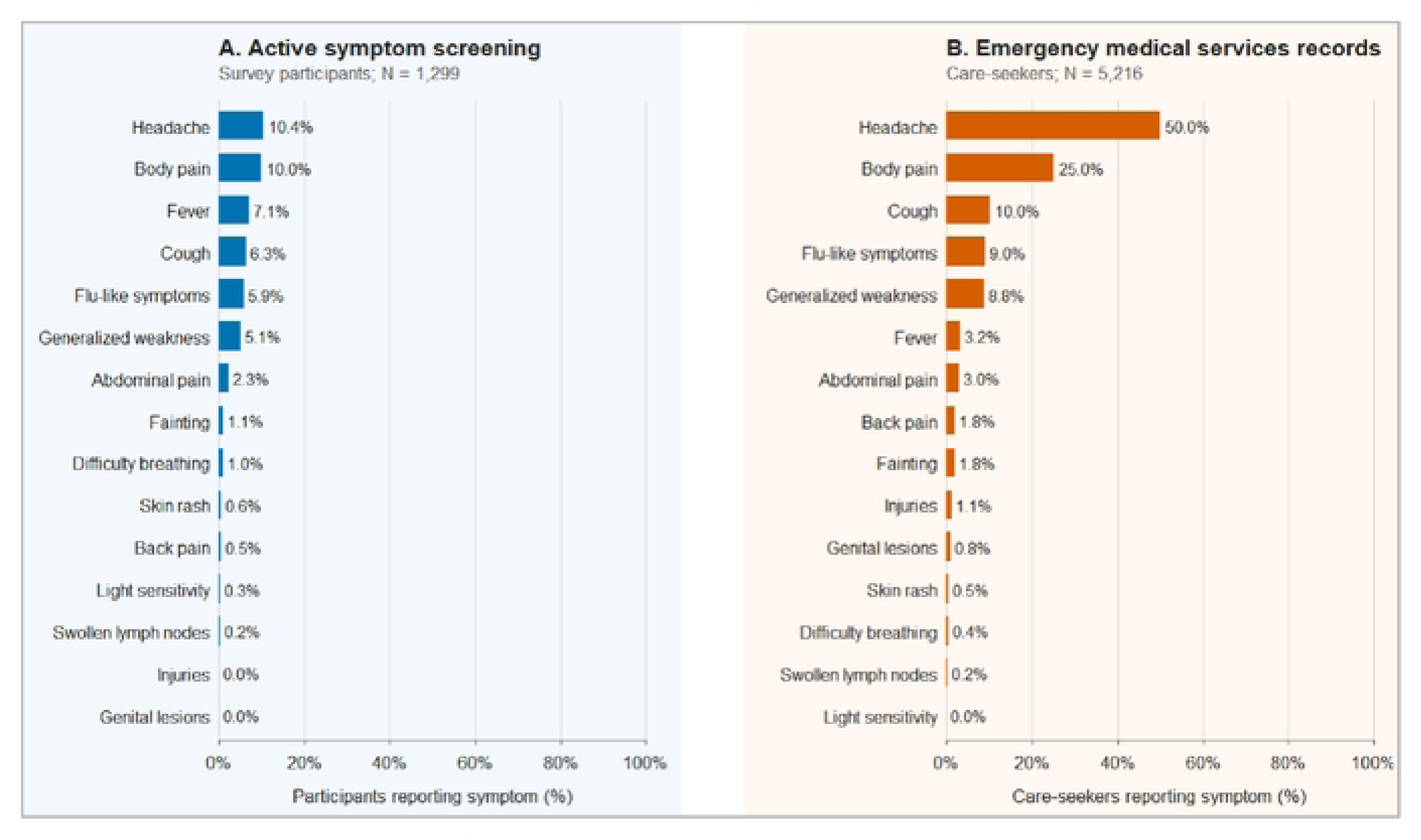
Symptoms identified through active screening (panel A) and emergency medical services records (panel B) during the Uganda Martyrs’ commemoration mass gathering, May 29–June 5, 2025.

### Mpox-related syndromic presentations in EMS records

Of the 5,216 pilgrims who sought medical care, 3,415 (65.5%) had at least one presenting sign or symptom documented in their EMS records. Among these records, 628 (18.4%) contained one or more symptoms or syndromic presentations included in the broader mpox-related symptom complex. However, rash and skin-lesion findings were not consistently documented; therefore, the complete mpox-compatible illness definition could not be applied to the EMS data. The 628 presentations consequently represent a broad, non-specific syndromic signal rather than suspected or confirmed mpox cases. Symptom patterns from active screening and EMS records are presented separately in Figure 2.

### Provisional diagnoses among pilgrims who sought medical care, 29 May–5 June 2025

Among 5,216 pilgrims who sought medical care during the Uganda Martyrs’ commemoration, the most frequently recorded provisional diagnoses were peptic ulcer disease (10.8%), respiratory tract infection (482; 9.2%), malaria (190; 3.6%), urinary tract infection (96; 1.8%), and gastritis (79; 1.5%). Less frequently recorded diagnoses included pharyngitis (43; 0.8%), conjunctivitis (38; 0.7%), and cellulitis or other skin infection (31; 0.6%) (Figure 3).

**Figure 3.**
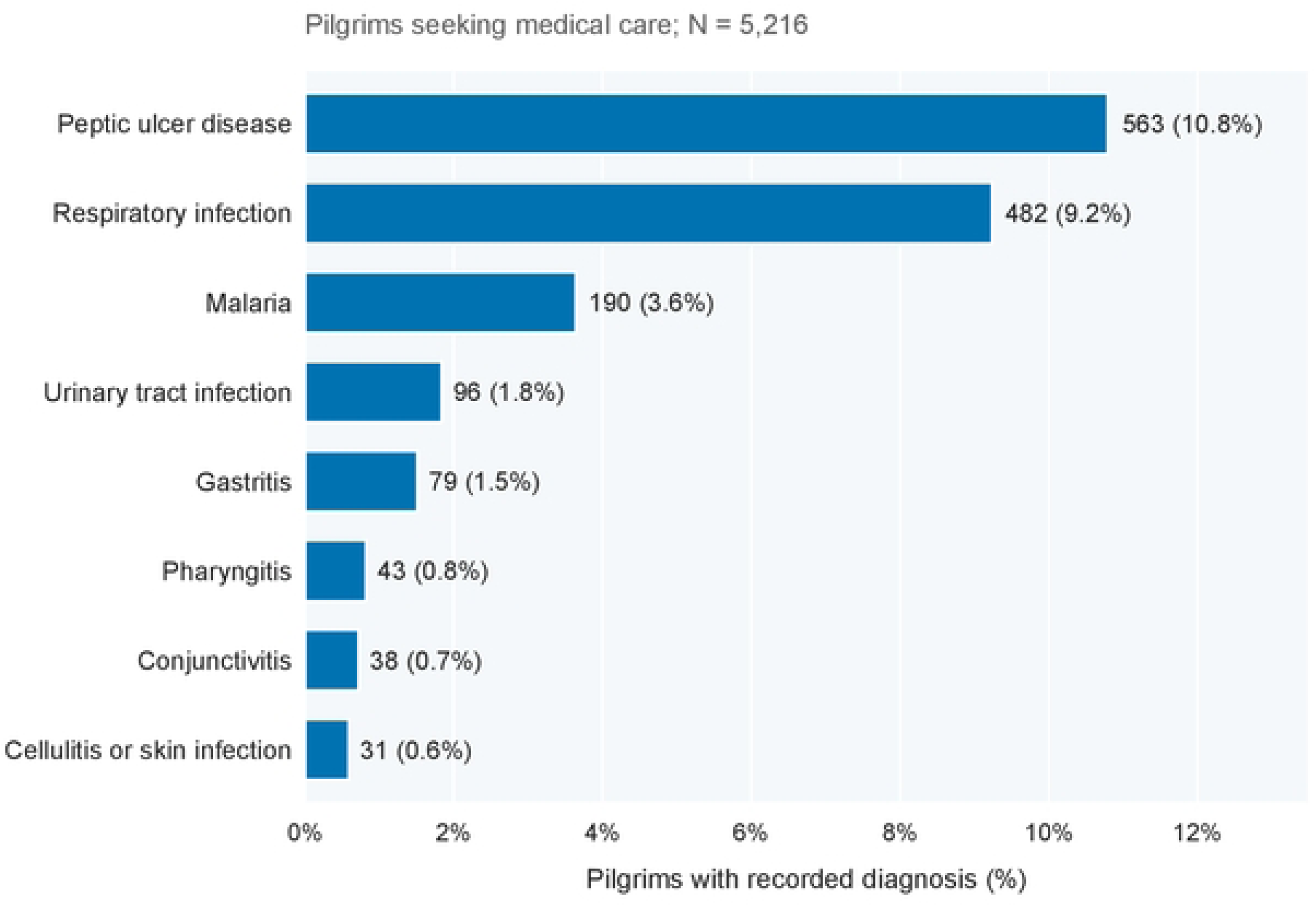
Common provisional diagnoses among pilgrims who sought medical care during the Uganda Martyrs’ commemoration mass gathering, May 29–June 5, 2025.

## Discussion

During the 2025 Uganda Martyrs’ Day commemoration, active symptom screening identified relatively few mpox-compatible presentations in the general pilgrim population, whereas emergency medical services records identified a stronger syndromic signal among pilgrims who sought care. This contrast suggests that future mass-gathering surveillance should combine broad active screening with intensified clinical assessment, rapid case investigation, and clear referral and testing pathways for symptomatic care-seekers.

The higher frequency of mpox-compatible syndromes among healthcare-seeking pilgrims than among the actively screened general pilgrim population is consistent with findings from syndromic surveillance during the 2022 Uganda Martyrs’ Day commemoration (8). That assessment similarly found that common acute illnesses predominated, while signals for priority epidemic-prone diseases were infrequent in the broader screened population. The present evaluation extends those findings by examining surveillance during an active national mpox outbreak and by directly comparing syndromic signals from population-based screening with those from a healthcare-seeking population.

The findings also highlight the inherent limitations of syndromic surveillance for mpox. Screening symptoms such as fever, headache, body aches, weakness, rash, and sore throat overlap with many common conditions encountered at mass gatherings, including malaria, respiratory and skin infections, dehydration, fatigue, and gastrointestinal illness (8, 15). Broad case definitions are valuable for early warning but may generate numerous false alerts unless they are linked to prompt clinical assessment and laboratory confirmation (8, 16). Operationally, surveillance systems may therefore need to balance sensitivity with specificity: they could minimize missed mpox cases without overwhelming field teams or misclassifying routine illnesses as outbreak-related events (14, 15).

The range of conditions managed by emergency medical services underscores the need for all-hazards preparedness at mass gatherings. Respiratory, febrile, gastrointestinal, urinary, ocular, throat, and skin conditions are expected where pilgrims experience crowding, prolonged travel, outdoor exposure, shared sanitation, and limited rest(15). Surveillance for mpox should therefore be integrated into a broader clinical and public health response capable of managing common illnesses while rapidly assessing, referring, and testing attendees with unusual or high-priority syndromes (6, 17).

For future religious mass gatherings held during active infectious disease transmission, syndromic surveillance should be linked to a predefined response pathway. Medical posts should use standardized symptom documentation and triage criteria for priority syndromes, with access to clinical assessment, specimen collection, referral, testing, treatment, and post-event follow-up (1, 12). Clear public messaging could help attendees recognize concerning symptoms and seek care promptly during and after the mass gathering (17, 18). These findings reinforce the need for integrated preparedness for recurring mass gatherings in Uganda. Active symptom screening and healthcare-based surveillance provide complementary information and should be embedded within a broader public health system that includes infection prevention and control, effective risk communication, laboratory support, and clear referral pathways. Preparedness plans should also ensure surge capacity for clinical assessment, specimen collection, testing, and public health response, while engaging religious leaders and community structures to promote timely reporting and appropriate preventive behaviours among pilgrims (15, 19).

### Limitations

This assessment has several limitations. First, mpox-compatible illness was identified using a syndromic case definition without laboratory confirmation, creating the potential for both false-positive and false-negative classification. Common illnesses such as malaria and other viral infections may have met the syndromic criteria, while mild or atypical mpox presentations could have been missed. Second, symptom information from the survey was self-reported and may have been affected by recall error. Third, emergency medical services records primarily documented provisional diagnoses rather than complete symptom profiles, which may have resulted in under-ascertainment of mpox-compatible presentations. Finally, although systematic sampling was used, crowd density, fluctuating pilgrim movement, and non-response during peak periods may have limited the representativeness of the surveyed population. The small number of participants meeting the mpox-compatible case definition also precluded analyses of factors associated with compatible illness.

Despite these limitations, this evaluation combined complementary community- and facility-based surveillance during an ongoing national mpox outbreak, providing a comprehensive assessment of syndromic surveillance in one of Africa’s largest annual religious mass gatherings. The findings offer practical evidence to inform surveillance and preparedness for future mass gatherings.

## Conclusion

During the 2025 Uganda Martyrs’ Day commemoration, active symptom screening identified few pilgrims with mpox-compatible illness, whereas medical-post surveillance identified more frequent presentations warranting clinical assessment. Future syndromic surveillance at mass gatherings should integrate structured active screening with real-time review of medical-post data and use predefined criteria for verifying and escalating priority infectious disease signals. Preparedness plans should assign responsibility for daily data review and specify pathways for clinical assessment, infection prevention, specimen collection, laboratory testing, notification, referral, and follow-up after attendees return home. After-action evaluations could assess data completeness, alert detection, laboratory turnaround, and response timeliness to strengthen surveillance at subsequent events.

## Data Availability

The datasets upon which our findings are based belong to the Uganda Public Health Fellowship Program. For confidentiality reasons, the data sets are not publicly available. The data sets can be availed upon reasonable request from the responsible officer with permission from the Uganda Public Health Fellowship Program. Request can be directed to

## Declarations

### Consent for publication

Not Applicable.

## Acknowledgements

We thank the pilgrims who participated in the survey for their time and willingness to share information during the 2025 Uganda Martyrs’ Day commemoration. We are also grateful to the community leaders at the Namugongo Catholic and Protestant shrines for their cooperation and for facilitating access to the shrines and study participants. We are grateful to the on-site medical teams from the Ministry of Health, Uganda Catholic Medical Bureau, and Uganda Protestant Medical Bureau for providing emergency medical services and for their support during data collection. We sincerely appreciate the surveillance officers from Makerere School of Public Health for their dedication in conducting the interviews and data extraction. We also thank the Ministry of Health for technical guidance and administrative clearance. Finally, we acknowledge the US Centers for Disease Control and Prevention (CDC) for supporting the Uganda Public Health Fellowship Program under which this assessment was conducted.

## Author contributions

JW, MDN, WN, AN and BL: Participated in the conception, design, analysis, and interpretation of the study and wrote the draft manuscript; RM and BK reviewed the report, reviewed the drafts of the manuscript for intellectual content and made multiple edits to the draft manuscript; RM, BK, LB, and ARA reviewed the manuscript to ensure intellectual content and scientific integrity. All authors read and approved the final manuscript.

## Author details

^1^Uganda Public Health Fellowship Program, Uganda National Institute of Public Health, Kampala, Uganda

^2^Department of Integrated Epidemiology, Surveillance and Public Health Emergencies, Ministry of Health, Kampala, Uganda

